# The interleukin-6/interleukin-23/Thelper-17-axis as a driver of neuro-immune toxicity in the major neurocognitive psychosis or deficit schizophrenia: a precision nomothetic psychiatry analysis

**DOI:** 10.1101/2022.02.23.22271393

**Authors:** Hussein Kadhem Al-Hakeim, Ali Fattah Alhusseini, Arafat Hussein Al-Dujaili, Monojit Debnath, Michael Maes

## Abstract

Schizophrenia and especially defcit schizophrenia (DSCZ)) are characterized by highly significantly increased activities of neuroimmunotoxic pathways and a generalized cognitive decline (G-CoDe). There are no data whether the interleukin(IL)-6/IL-23/Thelper-17 (IL6/IL23/Th17)-axis is more associated with DSCZ than with non-deficit schizophrenia (NDSCZ) and whether changes in this axis are associated with the G-CoDe and the phenome (a factor extracted from all symptom domains) of schizophrenia. This study included 45 DSCZ and 45 NDSCZ patients and 40 controls and delineated whether the IL6/IL23/Th17 axis, trace elements (copper, zinc) and ions (magnesium, calcium) are associated with DSCZ, the G-CoDe and phenome of schizophrenia. Increased plasma IL-23 and IL-6 levels were associated with Th17 upregulation, assessed as a latent vector (LV) extracted from IL-17, IL-21, IL-22, and TNF-α. The IL6/IL23/Th17-axis score, as assessed by a LV extracted from IL-23, IL-6, and the Th17 LV, was significantly higher in DSCZ than in NDSCZ and controls. We discovered that 70.7% of the variance in the phenome was explained by the IL6/IL23/Th17-axis (positively) and the G-CoDe and IL-10 (both inversely); and that 54.6% of the variance in the G-CoDe was explained by the IL6/IL23/Th17 scores (inversely) and magnesium, copper, calcium, and zinc (all positively). In conclusion, the pathogenic IL6/IL23/Th17-axis contributes to the generalized neurocognitive deficit and the phenome of schizophrenia and especially that of DSCZ due to its key role in peripheral inflammation and neuroinflammation and its consequent immunotoxic effects on neuronal circuits. These clinical impairments are more prominent in subjects with lowered IL-10, magnesium, calcium, and zinc.

## Introduction

Schizophrenia is a severe chronic neuropsychiatric disorder with a heterogeneous genetic and neurobiological background that affects about 0.75% of the world population (1). Recently, new qualitatively distinct phenotypes were discovered including the major neurocognitive psychosis (MNP, formerly called deficit schizophrenia) versus simple neurocognitive psychosis (SNP), non-remitters to treatment (NRTT) versus partial remitters to treatment (PRTT), and first-episode schizophrenia (FES) versus multiple-episode schizophrenia (MES) both with and without worsening (2). Around one-quarter of individuals with schizophrenia exhibit MNP which is characterized by more extreme neurocognitive deficits and symptomatology (3, 4).

MNP patients show more severe neurocognitive impairments than SNP patients in almost all cognitive domains including in sustained visual attention, working memory, strategy use, rule acquisition, attention set-switching, emotional recognition, semantic memory, episodic memory, and delayed recall, and recognition (5). Moreover, one common core (or latent construct) may be extracted from all these cognitive dysfunctions, named generalized cognitive decline (G-CoDe) (2, 5). MNP is characterized by intertwined increases in the severity of various symptom domains including psychosis, hostility, excitation, mannerism, and negative (PHEMN) symptoms, psychomotor retardation (PMR), and formal thought disorders (FTD) (6-8). Again, a common core underpins all seven symptom domains, conceptualized as the phenome of schizophrenia (9).

There is now evidence that schizophrenia, MNP, and FEP/FES are neuroimmune disorders. The first comprehensive theory of schizophrenia, known as the macrophage-T-lymphocyte theory was published in the 1990s (10). This theory considered that early neurodevelopmental disorders, as a consequence of intra-uterine infections, may predispose individuals to later injuries causing activation of immune-inflammatory and nitro-oxidative stress pathways. Since then, a growing amount of data have shown that immune-inflammatory processes in the peripheral blood and central nervous system (CNS) have a role in the onset of schizophrenia, primarily through systemic inflammation translating into microglial activation (11-19). Moreover, schizophrenia is not only characterized by activation of the immune-inflammatory response system (IRS), due to activated M1 macrophages, as well as T helper (Th)1 and Th17 cells but also by activation of the compensatory immune-regulatory system (CIRS) which tends to downregulate the primary IRS thereby preventing hyperinflammation (20). Key players in the CIRS are negative immunoregulatory cytokines produced by Th2 (IL-4) and T regulatory (Treg) (IL-10) cells (16). Importantly, relative deficits in the CIRS are associated with a worse outcome of FEP (21) and with increased neurocognitive impairments and the worsening in FEP and MEP (22). Moreover, increased neurotoxicity due to increased levels of toxic cytokines (e.g., IL-1β, IL-6, IL-8, TNF-α, and IFN-γ) and chemokines (e.g. CCL11, CCL2, CXCL8, and CXCL10) is signicantly associated with the G-CoDe and the phenome of schizophrenia (5). MNP is characterized by highly significant increases in neuroimmunotoxic pathways, attenuated CIRS protection, as well as increased phenome and G-CoDe scores (23-25).

Increasing evidence now suggests that Th17 cells are involved in the immunopathogenesis of schizophrenia (26) and a possible role of the IL-23/IL-17 pathway has been described (27-29). In addition, proinflammatory cytokines produced by pathogenic Th17 cells, such as IL-17 and IL-21 (30), have a significant role in the onset and progression of schizophrenia (26, 31, 32). Mediators produced by macrophages and dendritic cells such as IL-23, IL-1β, and IL-6 mediate the production of Th1, Th2, and Th17 cytokines (33, 34). IL-23, an IL-12 family proinflammatory cytokine, is vital in the IL-17-mediated immune response and in the survival and growth of pathogenic Th17 cells (35). IL-23 together with IL-6 and IL-1β or TGF-β stimulates naïve T cells to acquire the pathogenic Th17 phenotype (producing IL-17, IL-21, IL-22, and GM-CSF), whereas without IL-23 (stimulated by IL-6 and TGF-β) more homeostatic or non-pathogenic Th17 cells (producing IL-17, IL-21 and IL-10) are acquired (36-38). Increased production of IL-23 also drives TNF-α which together with IL-23 and IL-17 shapes the TNF/IL-23/IL-17 axis (39). The latter plays a key role in inflammation and (auto)immune disorders including rheumatoid arthritis, inflammatory bowel disease, multiple sclerosis, and type 1 diabetes mellitus (37). In addition, also IL-6 induces the production of IL-17 (40) and IL-21 in activated T cells (30). IL-21, in turn, induces the differentiation of Th17 cells in the presence of TGF-β1, while Th17 cells produce IL-21, which plays an autocrine role in Th17 cell development (41, 42).

There are also some reports that trace elements such as copper and zinc and ions such as calcium and magnesium as well as serum albumin may play a role in schizophrenia. Thus, serum copper is elevated in schizophrenia patients (43), while lower zinc is shown in a recent meta-analysis (44). Patients with schizophrenia have often lowered serum and intracellular levels of magnesium, while magnesium treatment may improve psychosis (45). There is also some evidence that aberrations in calcium signaling are associated with neurocognitive impairments (46) and that lowered albumin in may contribute to the severity of the phenome of schizophrenia (47).

Nevertheless, there are no data whether the IL-6, IL-23, and Th17 (IL-17, TNF-α, IL-21 and IL-22) axis is more associated with MNP than with SNP and whether changes in this axis are associated with the G-CoDe and the phenome of schizophrenia. Hence, this study was conducted to delineate a) whether the IL-6/IL-23/Th17 axis is activated in MNP and is associated with impairments in the G-CoDe and the phenome; b) the associations between IL-6 and IL-23 and Th17 effector cytokines; and c) the role of IL-10, copper, zinc, calcium and magnesium in the neuro-immune pathophysiology of MNP.

## Subjects and Methods

### Participants

Ninety patients with schizophrenia and forty healthy controls were included in this study. Patients were recruited between February and June 2021 at “The Psychiatry Unit,” Al-Hakeem General Hospital, Najaf Governorate, Iraq. All patients were stable for at least 12 weeks. Family and friends of personnel or friends of patients served as controls. Patients and controls were excluded if they had ever taken immunosuppressive treatments or glucocorticoids, or if they had been diagnosed with a neurodegenerative or neuroinflammatory illness such as Alzheimer’s disease, Parkinson’s disease, multiple sclerosis, or stroke. Additionally, individuals with (auto)immune diseases such as inflammatory bowel disease, rheumatoid arthritis, COPD, psoriasis, or diabetes mellitus were excluded. None of the controls had a present or lifetime DSM-IV-TR axis I diagnosis or a family history of schizophrenia or psychosis. Patients having axis-1 DSM-IV-TR disorders such as bipolar disorder, major depression, schizoaffective disorder, obsessive-compulsive disorder, psycho-organic illnesses, or substance use disorders were excluded. CRP levels were < 6 mg/L in all subjects, indicating that there was no overt inflammation.

The study followed Iraqi and international privacy and ethics laws. Before participating in this study, all participants and first-degree relatives of participants with schizophrenia gave written informed consent (legal representatives are mother, father, brother, spouse, or son). The study was approved by the ethics committee (IRB) of the College of Science, University of Kufa, Iraq (82/2020), which follows the Declaration of Helsinki’s International Guideline for Human Research Protection.

### Clinical assessments

To collect patient and control data, a senior psychiatrist with expertise in schizophrenia conducted semi-structured interviews. The Mini-International Neuropsychiatric Interview (M.I.N.I.) was used to diagnose schizophrenia using DSM-TR criteria. The same psychiatrist evaluated the SDS (48) for the diagnosis of primary deficit schizophrenia, as well as the Positive and Negative Syndrome Scale (PANSS), the Scale for the Assessments of Negative Symptoms (SANS) (49), the Brief Psychiatric Rating Scale (BPRS) (50), and the Hamilton Depression (HAM-D) and Anxiety (HAM-A) rating scales (51, 52). We calculated z-unit weighted composite scores reflecting psychosis, hostility, excitation, mannerism, PMR and FTD symptom domains as previously reported using BPRS, HAM-D, HAM-A, and PANSS items and the total SANS score was used to assess negative symptoms (8, 53-55). Based on the SANS score and the 6 computed z composite scores reflecting PHEM, PMR, and FTD we then extracted the first factor from these data, named the “phenome”. On the same day, a research psychologist performed neuropsychological probes using the Brief Assessment of Cognition in Schizophrenia (BACS) (56) while remaining blind to the clinical diagnosis. The latter battery consists of the Digit Sequencing Task (which measures working memory), the List Learning test (which tests verbal episodic memory), Controlled Word Association (which assesses letter fluency), Category Instances (which measures semantic fluency), the Tower of London (to probe executive functions), Symbol Coding (which probes attention), and the token motor task. The total score was computed. Based on our previous findings (5) that one latent vector underpins all cognitive tests, we here extract the first factor from the 7 BACS test, and named this factor the “G-CoDe” (generalized cognitive decline). Tobacco use disorder (TUD) was diagnosed in accordance with DSM-IV-TR criteria. The following formula was used to compute the body mass index (BMI): body weight (kg) / length (m^2^).

### Assays

Each subject’s fasting venous blood was obtained in the early morning hours. After 15 minutes at room temperature, blood was allowed to coagulate for 10 minutes before being centrifuged at 3000 rpm for 10 minutes. Separated serum was then transferred to Eppendorf tubes and stored at -80 °C until analysis. The concentrations of CRP in serum were measured using a kit supplied by Spinreact^®^ (Barcelona, Spain). The test is based on the notion of latex agglutination. Serum albumin, calcium, and magnesium concentrations were determined using spectrophotometric kits provided by Biolabo^®^ (Maizy, France). Copper was determined spectrophotometrically by kits supplied by LTA Co., (Milano, Italy). Zinc concentrations were determined using a kit provided by Cenrionic GmbH (Wartenberg, Germany). A panel of cytokines that serve as positive regulators (IL-1β, IL-6, IL-23), negative regulators (IL-10), mediators (IL-21) and effectors (IL-17, IL-22, TNF-α, G-CSF) of the IL6/IL23/Th17 pathway was considered to obtain a clear insight into the role of Th17-pathway in MNP. Serum IL-6, IL-10, G-CSF, and IL-1β were measured using commercial ELISA sandwich kits provided by Sunlong Biotech Co., Ltd (Zhejiang, China), and Melsin Medical Co. (Jilin, China) provided the other ELISA kits (IL-17, IL-21, IL-22, IL-23, and TNF-α). The intra-assay coefficients of variation (CV) for all the assays were <10.0% (precision within-assay). The sensitivities of the kits were 1.0 pg/ml for IL-10, IL-17, IL-21, IL-22, IL-23, TNF-α, G-CSF, and 0.1 pg/ml for IL-1β and IL-6.

### Statistical analysis

To compare scale variables between groups, one-way analysis of variance was employed, while analysis of contingency tables (two tests) was used to analyze category variables. We used multivariate GLM analysis to examine the association between the 14 biomarkers and the diagnosis while entering age, gender, education, BMI, TUD, and drug use as covariates. The between-subject effects tests were employed to investigate the impact of significant explanatory factors on each of the observed biomarkers. As a result, we calculated estimated marginal means (SE) values produced by the model (adjusted for the significant confounders). These multiple associations were subjected to p-correction for false discovery rate (FDR) (57) and the protected Least Significant Difference (LSD) tests were used to assess multiple pair-wise differences. Multiple regression analysis (automated approach with p-to-entry of 0.05 and p-to-remove of 0.06 while assessing the change in R^2^) was used to determine the important biomarkers that predict the phenome in schizophrenia and all participants combined. Multivariate normality (Cook’s distance and leverage), multicollinearity (using tolerance and VIF), and homoscedasticity (using White and modified Breusch-Pagan tests for homoscedasticity) were checked. These regression analyses’ results were always bootstrapped using 5.000 bootstrap samples, and the latter results are shown if the results were not concordant. We used two-step cluster analysis to derive meaningful clusters from the data set. Feature reduction was performed using exploratory factor analysis. IBM SPSS Windows version 25, 2017 was used to conduct the statistical analysis.

Partial Least Squares (PLS)-SEM pathway analysis is a statistical approach for predicting complex cause-effect linkages utilizing both discrete indicators (variables) and latent variables (factors based on a collection of highly connected indicators) (58). Without imposing distributional assumptions on the data, PLS allows estimation of complex multi-step mediation models with several latent constructs, single indicator variables, and structural pathways (associations between indicators or latent vectors). This strategy has recently been used to develop novel precision nomothetic models of affective disorders and schizophrenia by combining the many components of a disease into a causal-effect, mediation model (9, 23). A causative framework is developed utilizing causome, protectome, adverse outcome pathways, and phenome indicators (either single indicators or latent vectors), which is then analyzed and cross-validated using PLS pathway analysis on bootstrapped data (e.g. 5.000 samples). As a result, pathway coefficients (with exact p values), specific indirect (mediated) and total effects are estimated to determine the influence of direct and mediated pathways. According to the power analysis, the predicted sample size for a multiple regression analysis (which is applicable to PLS) should be at least 70 to obtain a power of 0.8, an effect size of 0.2, and an alpha of 0.05 with 5 preset variables. Complete path analysis with 5.000 bootstrap samples was conducted only when the outer and inner models matched the following quality criteria: a) model quality as measured by the SRMR index is less than 0.08; b) outer model loadings on the latent vectors are greater than 0.666 at p < 0.001; and c) the latent vectors exhibit accurate construct validity as measured by average variance extracted > 0.5, Cronbach’s alpha > 0.7, rho A > 0.7, and composite reliability > 0.8. Compositional invariance was investigated using Predicted-Oriented Segmentation analysis, Multi-Group Analysis, and Measurement Invariance Assessment. The predictive performance of the model was evaluated using PLSpredict with a 10-fold cross-validation.

## Results

### Sociodemographic data

**Table 1** shows the sociodemographic and clinical data of MNP versus SNP and controls. There were no significant differences in age, sex, BMI, marital status, TUD, residency, and employment status between MNP and SNP and healthy controls. Both patients groups were somewhat less educated than the control group, while there were no significant differences in education level between both patient groups. MNP patients have a significantly higher (p<0.001) positive family history of schizophrenia than the SNP group. Since part of the patients were treated with olanzapine (n=62), clozapine (n=12), fluphenazine (n=27), haloperidol (n=7), quietapine (n=5), risperidal (n=16), benzodiazepines (n=19), and mood stabilizers (n=39) we have examined the impact of these drugs on the clinical scores. After adjusting for age, sex, BMI, smoking, education, and the drug state of the subjects, the major symptoms domain scores (psychosis, hostility, excitement, mannerism, SANS, PMR, and FTD) were significantly different between the three study samples and increased from controls → SNP → MNP. These differences remained significant after FDR p-correcion at p<0.001.

**Table 1:** Sociodemographic and clinical data of healthy controls (HC) and schizophrenia patients divided into those with major neurocognitive (MNP) versus simple neurocognitive (SNP) psychosis

| Variables | HC <sup>A</sup><br>(n=40) | SNP <sup>B</sup><br>(n=45) | MNP <sup>C</sup><br>(n=45) | F/$\chi^2$ | df | p |
| --- | --- | --- | --- | --- | --- | --- |
| Age (years) | 36.8 (12.0) | 37.4 (12.6) | 37.5 (12.3) | 0.04 | 2/127 | 0.965 |
| Sex (Female/Male) | 20/20 | 18/27 | 19/26 | 0.93 | 2 | 0.627 |
| Single/married | 17/23 | 19/26 | 19/26 | 0.001 | 2 | 1.000 |
| BMI (kg/m<sup>2</sup>) | 26.87 (3.57) | 26.86 (3.92) | 27.17 (3.81) | 0.097 | 2/127 | 0.908 |
| TUD (No/Yes) | 24/16 | 26/19 | 27/18 | 0.06 | 2 | 0.970 |
| Residency Urban/Rural | 16/24 | 17/28 | 17/28 | 0.058 | 2 | 0.972 |
| Employment (No/Yes) | 14/26 | 16/29 | 20/23 | 5.33 | 2 | 0.255 |
| Education (years) | 12.1 (3.3) <sup>B,C</sup> | 9.2 (3.6) <sup>A</sup> | 9.5 (3.5) <sup>A</sup> | 8.72 | 2/127 | <0.001 |
| Age at onset (years) | - | 28.4 (9.0) | 26.0 (12.8) | 1.141 | 1/88 | 0.288 |
| Family history (No/Yes) | 40/0 <sup>B,C</sup> | 29/16 <sup>A,B</sup> | 23/22 <sup>A,C</sup> | 25.80 | 2 | <0.001 |
| Psychosis (z scores) | -1.000 (0.184) <sup>B,C</sup> | -0.234(0.113) <sup>A,B</sup> | 1.148(0.088) <sup>A,C</sup> | 157.45 | 2/115 | <0.001 |
| Hostility (z scores) | -0.908 (0.243) <sup>B,C</sup> | -0.165(0.149) <sup>A,B</sup> | 1.157(0.116) <sup>A,C</sup> | 82.78 | 2/115 | <0.001 |
| Excitement (z scores) | -1.337 (0.265) <sup>B,C</sup> | -0.183(0.163) <sup>A,B</sup> | 0.885(0.127) <sup>A,C</sup> | 63.70 | 2/115 | <0.001 |
| Mannerism (z scores) | -2.006 (0.496) <sup>B,C</sup> | -0.035(0.305) <sup>A,B</sup> | 2.245(0.237) <sup>A,C</sup> | 71.86 | 2/115 | <0.001 |
| PMR (z scores) | -0.885 (0.215) <sup>B,C</sup> | .034(0.132) <sup>A,B</sup> | 1.258(0.103) <sup>A,C</sup> | 102.25 | 2/115 | <0.001 |
| FTD (z scores) | -1.076 (0.235) <sup>B,C</sup> | -0.316(0.144) <sup>A,B</sup> | 1.084(0.112) <sup>A,C</sup> | 98.33 | 2/115 | <0.001 |
| Total SANS (z scores) | -1.173( 0.202) <sup>B,C</sup> | -0.008(0.124) <sup>A,B</sup> | 1.047(0.097) <sup>A,C</sup> | 108.71 | 2/115 | <0.001 |

Results are shown as mean (SD), except the symptom domain scores which are shown as estimated marginal mean (SE) values after adjusting for the effects of age, sex, education, BMI (body mass index), smoking and the drug state. <sup>A,B,C</sup>: pairwise comparisons between group means. FTD: Formal thought disorder, PMR: Psychomotor retardation, SANS: Scale for the Assessment of Negative Symptoms, TUD: Tobacco use disorder.

### Differences in biomarkers between the study groups

Multivariate GLM analysis (**Table 2)** was used to assess the relationships between biomarkers and diagnosis after controlling for age, BMI, sex, smoking and the drug state. There were highly significant differences in the biomarkers across the groups (which remained significant after FDR p-correction at p<0.012), but no significant impact of the covariates.Tests for between-subject effects in **Table 3**, which shows the estimated marginal means, revealed that IL-23, IL-6, IL-17, and TNF-α were significantly higher in MNP than in SNP and controls and that IL-17 and TNF-α were higher in SNP than in controls. IL-1β, IL-22, G-CSF, IL-21, and IL-10 were significantly higher in schizophrenia than in controls. There were no differences in albumin between the study groups and zinc levels were lowest in both schizophrenia groups. Magnesium was significantly lowered in MNP as compared with the other two groups, whereas calcium was lower in MNP than in SNP. We found no significant effects of the drug state on the single biomarkers (tested with multivariate GLM analysis and tests for between-subjects effects) even without p correction for FDR.

**Table 2.** Results of multivariate GLM analysis examining the associations between immune markers and diagnosis, namely healthy controls and schizophrenia paients divided into those with major (MNP) and simple (SNP) neurocognitive psychosis.

| Tests | Dependent variables | Explanatory variables | F | df | p |
| --- | --- | --- | --- | --- | --- |
| **Multivariate** | All 14 biomarkers | Diagnosis | 5.22 | 28/208 | <0.001 |
|  |  | Sex | 1.02 | 14/103 | 0.437 |
|  |  | Age | 1.35 | 14/103 | 0.193 |
|  |  | BMI | 0.84 | 14/103 | 0.62 |
|  |  | Smoking | 0.42 | 14/103 | 0.964 |
| **Between-subject effects** | Albumin | Diagnosis | 0.09 | 2/116 | 0.916 |
|  | Magnesium | Diagnosis | 6.54 | 2/116 | 0.002 |
|  | Calcium | Diagnosis | 2.83 | 2/116 | 0.063 |
|  | Copper | Diagnosis | 4.76 | 2/116 | 0.010 |
|  | Zinc | Diagnosis | 6.04 | 2/116 | 0.003 |
| | Interleukin (IL)-1β | Diagnosis | 9.65 | 2/116 | <0.001 |
|  | IL-23 | Diagnosis | 6.74 | 2/116 | 0.002 |
|  | IL-22 | Diagnosis | 9.39 | 2/116 | <0.001 |
|  | IL-6 | Diagnosis | 5.48 | 2/116 | 0.005 |
|  | G-CSF | Diagnosis | 2.90 | 2/116 | 0.059 |
|  | IL-21 | Diagnosis | 5.94 | 2/116 | 0.004 |
|  | IL-17 | Diagnosis | 28.27 | 2/116 | <0.001 |
|  | IL-10 | Diagnosis | 6.05 | 2/116 | 0.003 |
| | TNF-α | Diagnosis | 16.75 | 2/116 | <0.001 |
| **Univariate** | IL6/IL23/Th17 axis score | Diagnosis | 94.09 | 2/123 | <0.001 |

The results of these GLM analyses were adjusted for use of olanzapine, clozapine, modicate, kemadrine, risperidone, benzodiazepines, mood stabilizers (not shown as all non-significant).
G-CSF: Granulocyte colony stimulating factor, TNF-α : tumor necrosis factor, IL6/IL23/Th17: a latent vector score extracted from 6 cytokines, namely IL-6, IL-23, IL-17, IL-21, IL-22, and TNF-α .

**Table 3.** Model-generated estimated marginal means of the different immune markers in healthy controls (HC) and patients with schizophrenia divided into those with major (MNP) and simple (SNP) neurocognitive psychosis

| Variables | HC <sup>A</sup> | SNP <sup>B</sup> | MNP <sup>C</sup> |
| --- | --- | --- | --- |
| Albumin g/l | 46.93 (3.44) | 46.33 (2.14) | 45.85 (1.66) |
| Magnesium mM | 1.008 (0.060) <sup>C</sup> | 1.019 (0.037) <sup>C</sup> | 0.911 (0.029) <sup>A,B</sup> |
| Calcium mM | 2.377 (0.122) | 2.367 (0.076) <sup>C</sup> | 2.227 (0.059) <sup>B</sup> |
| Copper mg/l | 0.785 (0.119) | 0.728 (0.074) <sup>C</sup> | 0.914 (0.057) <sup>B</sup> |
| Zinc mg/l | 0.668 (0.075) <sup>B,C</sup> | 0.503 (0.047) <sup>A,C</sup> | 0.603(0.036) <sup>A,B</sup> |
| Interleukin-(IL)-1β pg/ml | 2.5 (0.4) <sup>B,C</sup> | 3.6 (0.3) <sup>A</sup> | 4.0 (0.2) <sup>A</sup> |
| IL-23 pg/ml | 15.9 (6.8) <sup>C</sup> | 17.6 (4.2) <sup>C</sup> | 29.4 (3.3) <sup>A,B</sup> |
| IL-22 pg/ml | 15.8 (3.4) <sup>B,C</sup> | 25.9 (2.1) <sup>A</sup> | 27.2 (1.6) <sup>A</sup> |
| IL-6 pg/ml | 5.9 (1.6) <sup>C</sup> | 5.8 (1.0) <sup>C</sup> | 8.5 (0.8) <sup>A,B</sup> |
| G-CSF pg/ml | 77.6 (21.2) <sup>B,C</sup> | 109.8 (13.1) <sup>A</sup> | 118.2 (10.2) <sup>A</sup> |
| IL-21 pg/ml | 170.7 (41.9) <sup>B,C</sup> | 255.0 (26.0) <sup>A</sup> | 285.8 (20.2) <sup>A</sup> |
| IL-17 pg/ml | 23.3 (4.5) <sup>B,C</sup> | 43.5 (2.8) <sup>A,C</sup> | 50.2 (2.2) <sup>A,B</sup> |
| IL-10 pg/ml | 6.0 (1.8) <sup>B,C</sup> | 10.5 (1.1) <sup>A</sup> | 10.7 (0.9) <sup>A</sup> |
| TNF-α pg/ml | 18.5 (5.7) <sup>B,C</sup> | 36.7 (3.5) <sup>A,C</sup> | 44.7 (2.8) <sup>A,B</sup> |
| IL23/IL6/Th17 z score | -1.051 (0.102) | 0.067 (0.096) | 0.867 (0.096) |

<sup>A,B,C</sup>: pairwise comparisons between group means. G-CSF: Granulocyte colony stimulating factor, TNF-α : tumor necrosis factor, IL23/IL6/Th17: a latent vector score extracted from 6 cytokines, namely IL-23, IL-6, IL-17, IL-21, IL-22, and TNF-α .

### Results of PLS analysis

Figure 1. shows the results of PLS path analysis with IL-23 and IL-6 as primary input variables and latent vectors extracted from all symptom domains (reflecting the phenome) and cognitive tests (reflecting G-CoDe) as output variables. In this model, IL-23 and IL-6 were considered to be related with a latent vector (LV) extracted from IL-17, IL-21, IL-22, and TNF-α (labeled Th17 LV) while the latter predicted G-CSF and IL-10. After feature selection, multi-group analysis, PLS predict analysis, and prediction-oriented segmentation, **Figure 1** shows the results of the PLS path analysis conducted on 5.000 bootstrap samples. The final PLS model did not contain non-significant pathways and variables. With SRMR=0.039, this model’s overall fit was more than sufficient. Additionally, the reliability of the factor constructs was satisfactory for all three factors, with Cronbach alpha > 0.754, rho A > 0.785, composite reliability > 0.845, and AVE > 0.58. At p <0.0001, all loadings on the outer models were more than 0.620. We discovered that 70.7% of the variance in the phenome could be explained by regression on the cognitome, Th17 LV (both positively) and IL-10 (inversely), and that 68.9% of the variance in the cognitome could be explained by education (positively) and the combination of IL-23 and Th17 LV. IL-23 and IL-6 explained 17.7% of the variance in the Th17 LV. The cognitome (0.558), the Th17 LV (0.093), and the phenome (0.624) all had acceptable construct cross-validated redundancies (as determined by PLS blindfolding). CTA demonstrated that the latent vectors were not conceptualized incorrectly as reflective models. PLS predict revealed that the indicators of the endogenous constructs had positive Q^2^ predict values, indicating that they outperformed the naïve benchmark. There were significant specific indirect effects of IL-23 on the phenome through the Th17 LV (t=2.82, p=0.002), the cognitome (t=1.68, p=0.047), the path from Th17 LV to the cognitome (t=3.64, p<0.001), and the path from Th17 LV to IL-10 (t=-1.84, p=0.033). Although the direct pathway from IL-6 to the phenome was not significant, the overall impact of IL-6 on the phenome was significant (t=1.74, p=0.041). Nonetheless, IL-23 (t=6.76, p<0.001) and the Th17 LV (t=5.10, p<0.001) had a much greater influence on the phenome.

**Figure 1.**
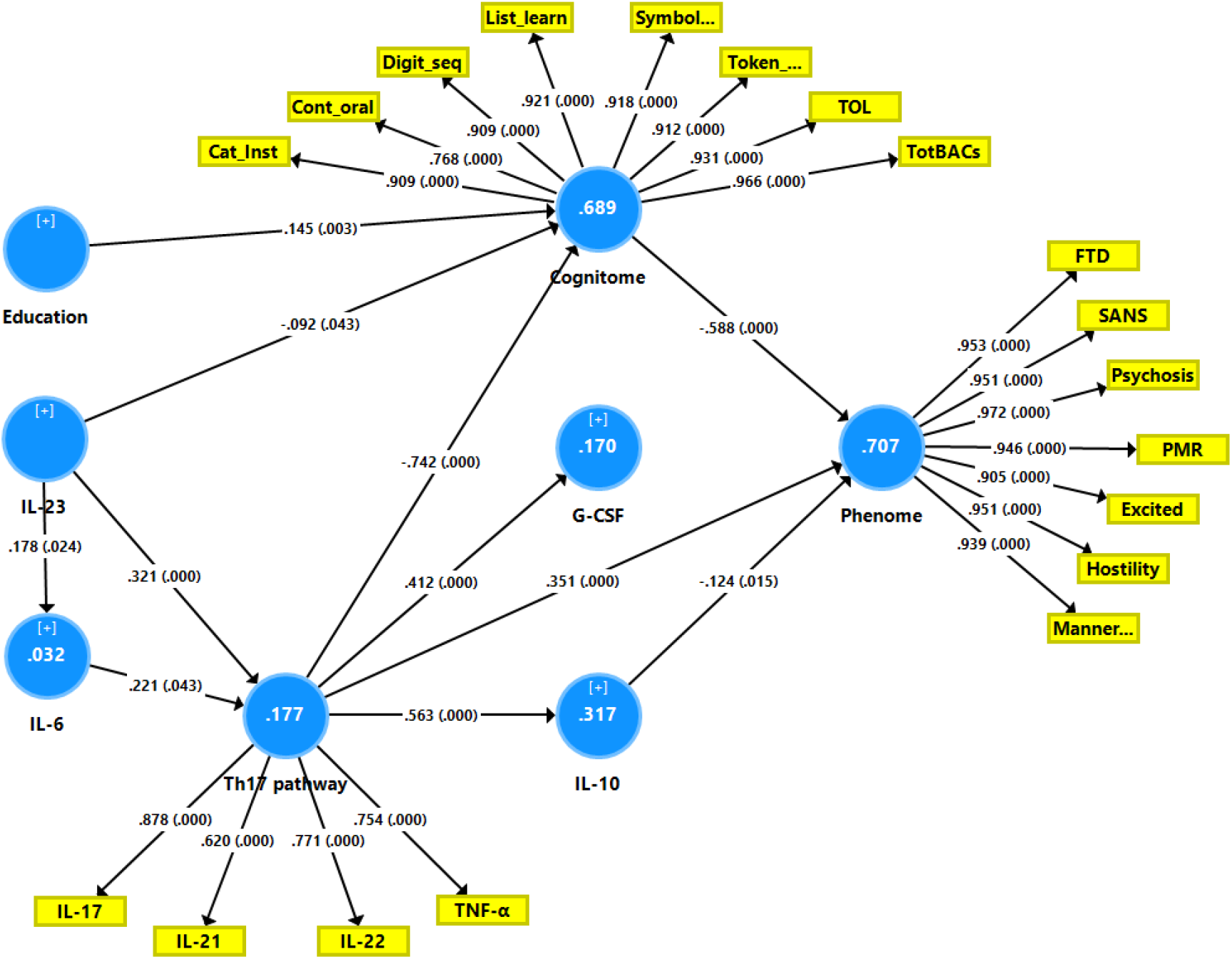
Results of Partial least Squares (PLS) path analysis. Interleukin (IL)-23 and IL-6 are primary input variables. Latent vectors extracted from T helper (Th)17 cytokines, including tumor necrosis factor (TNF)-α, and the generalized cognitive decline (G-CoDe) mediate the effects of IL-6 and IL-23 on the phenome of schizophrenia (outcome indicator). The latter is conceptualized as a latent vector extracted from all symptom domains (reflecting the phenome). The model also considers effects of Th-17 on granulocyte-colony stimulating factor (G-CSF) and IL-10. Cat_Inst: Category Instances; Cont_oral: Controlled Word Association; Digit_seq: Digit Sequencing Task List_learn: the List Learning test; Symbol..: Symbol Coding; Token..: token motor task; TOL: Tower of London FTD: formal thought disorders; SANS: negative symptoms; PMR: psychomotor retardation; Excited: excitation; Manner..: mannerism. White figures in circles: explained variance. Shown are the pathway coefficients (p values) and factor loadings (p values)

### Construction of pathway endophenotype and endophenotype classes

Based on the PLS findings that IL-6 and IL-23 are both associated with the Th17 LV we constructed a LV extracted from IL-6 and IL-23 (AVE=58.7%, composite reliability=0.738, loadings: 0.823 and 0.705, respectively) and then extracted a LV from this construct and the Th17 LV, yielding a new IL6/IL23/Th17 LV (AVE=70.8%, loadings: 0.841 and 0.841, respectively, composite reliability=0.844). Tables 2 and 3 show that this IL6/IL23/Th17 score was significantly different between the three study groups and increased from controls → MNP → SNP. In addition, we were able to extract a latent vector from the IL6/IL23/Th17 (loadings: 0.880), G-CoDe (−0.918) and phenome (0.918) scores (AVE=81.52%, composite reliability=0.912). Two step cluster analysis with the IL6/IL23/Th17, G-CoDe, and phenome entered as continuous variables and the diagnosis schizophrenia as categorical variable showed three clusters of patients with a silhouette measure of cohesion and separation of 0.66. The formed schizophrenia clusters exactly matched with the MNP (deficit) versus SNP (non-deficit) dichotomy. We found no significant effects of these drugs on the IL6/IL23/Th17 score (tested with univariate GLM analysis).

### Prediction of symptom domains and G-CoDe by the IL6/IL23/Th17 axis score

The first multiple regression analysis in **Table 4** entered the phenome LV as the dependent variable and the IL6/IL23/Th17 LV and G-CoDe scores as the explanatory variables, while allowing for the effects of sex, age, smoking, education, the trace elements, and ions. Table 4, regression #1 demonstrates that 68.8% of the variance in the phenome was explained by the regression on G-CoDe and IL6/IL23/Th17 scores. **Figures 2** and **3** show the partial regression plots of the phenome on both predictors (after considering the effects of the other covariate). In the restricted study sample of schizophrenia patients (regression #2) the same variables explained 28.5% of the variance in the phenome.

**Table 4.** Results of multiple regression analyses with the phenome of schizophrenia or the generalized cognitive decline (G-CoDe) score as dependent variables and biomarkers as explanatory variables.

| Dependent variables | Explanatory variables | $\beta$ | t | p | F<sub>model</sub> | df | p | R<sup>2</sup> |
| --- | --- | --- | --- | --- | --- | --- | --- | --- |
| #1. Phenome in all subjects | **Model** |  |  |  | **122.01** | **2/127** | **<0.001** | **0.658** |
|  | G-CoDe | -0.550 | -7.69 | <0.001 |  |  |  |  |
|  | IL6/IL23/Th17 axis | 0.328 | 4.60 | <0.001 |  |  |  |  |
| #2. Phenome in schizophrenia | **Model** |  |  |  | **17.33** | **2/87** | **<0.001** | **0.285** |
|  | G-CoDe | -0.312 | -3.23 | 0.002 |  |  |  |  |
|  | IL6/IL23/Th17 axis | 0.339 | 3.52 | <0.001 |  |  |  |  |
| #3. G-CoDe in all subjects | **Model** |  |  |  | **37.65** | **4/125** | **<0.001** | **0.546** |
|  | IL6/IL23/Th17 axis | -0.688 | -10.70 | <0.001 |  |  |  |  |
|  | Magnesium | 0.234 | 3.73 | <0.001 |  |  |  |  |
|  | Zinc | 0.126 | 2.05 | 0.043 |  |  |  |  |
|  | Copper | 0.143 | 2.19 | 0.030 |  |  |  |  |
| #43. G-CoDe in schizophrenia | **Model** |  |  |  | **11.40** | **3/86** | **<0.001** | **0.285** |
|  | IL6/IL23/Th17 axis | -0.282 | -3.06 | 0.003 |  |  |  |  |
|  | Magnesium | 0.366 | 3.99 | <0.001 |  |  |  |  |
|  | Calcium | 0.198 | 2.17 | 0.033 |  |  |  |  |

Phenome: conceptualized as the first principal component (PC) extracted from all symptom domains
G-CoDe: Index of generalized cognitive decline, conceptualized as the first PC extracted from all neurocognitive tests results
IL6/IL23/Th17: a latent vector score extracted from 6 cytokines, namely IL-6, IL-23, IL-17, IL-21, IL-22 and TNF-α .

**Figure 2.**
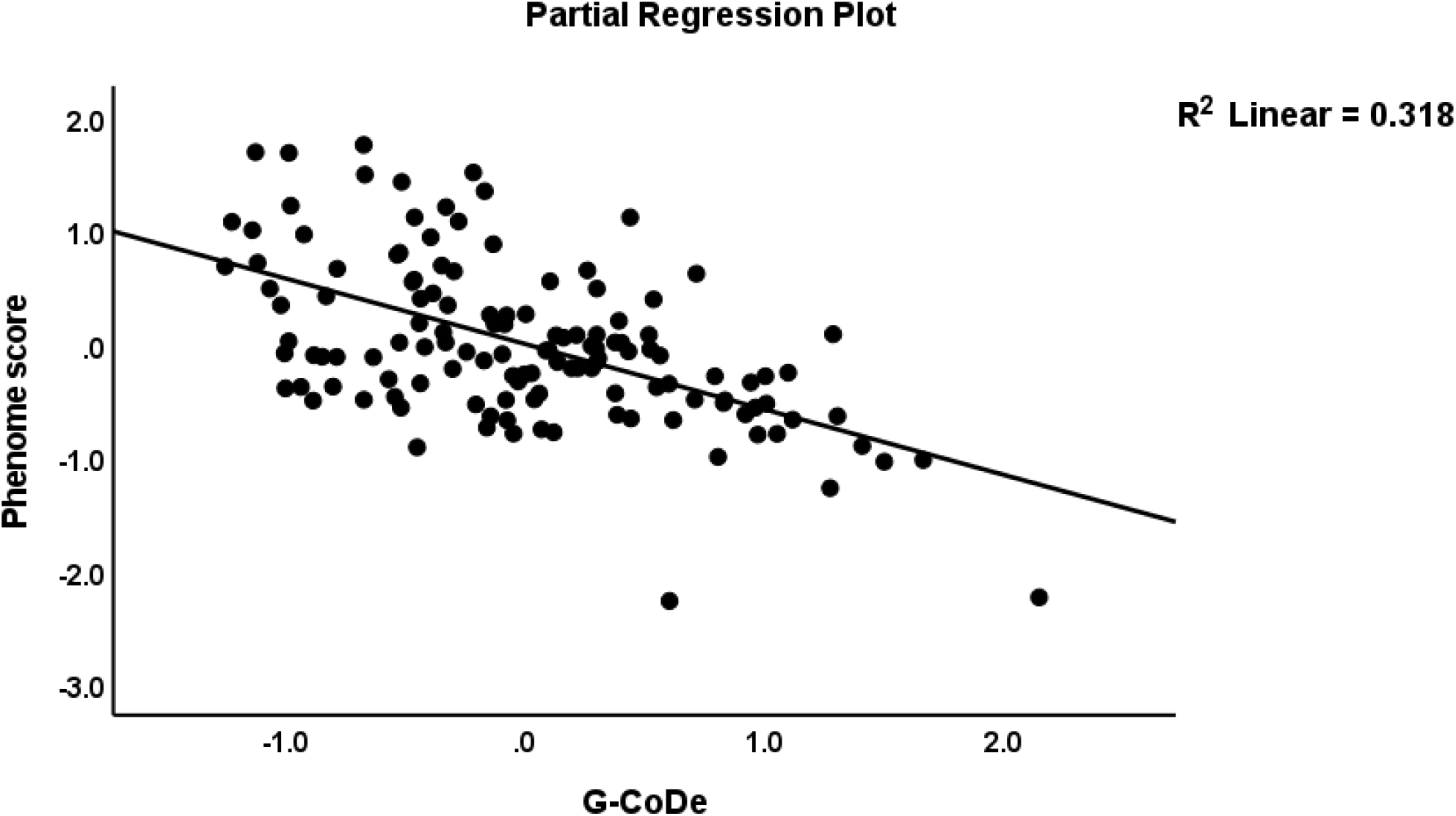
Partial regression plot of the phenome of schizophrenia on the generalized cognitive decline (G-CoDe) score

**Figure 3.**
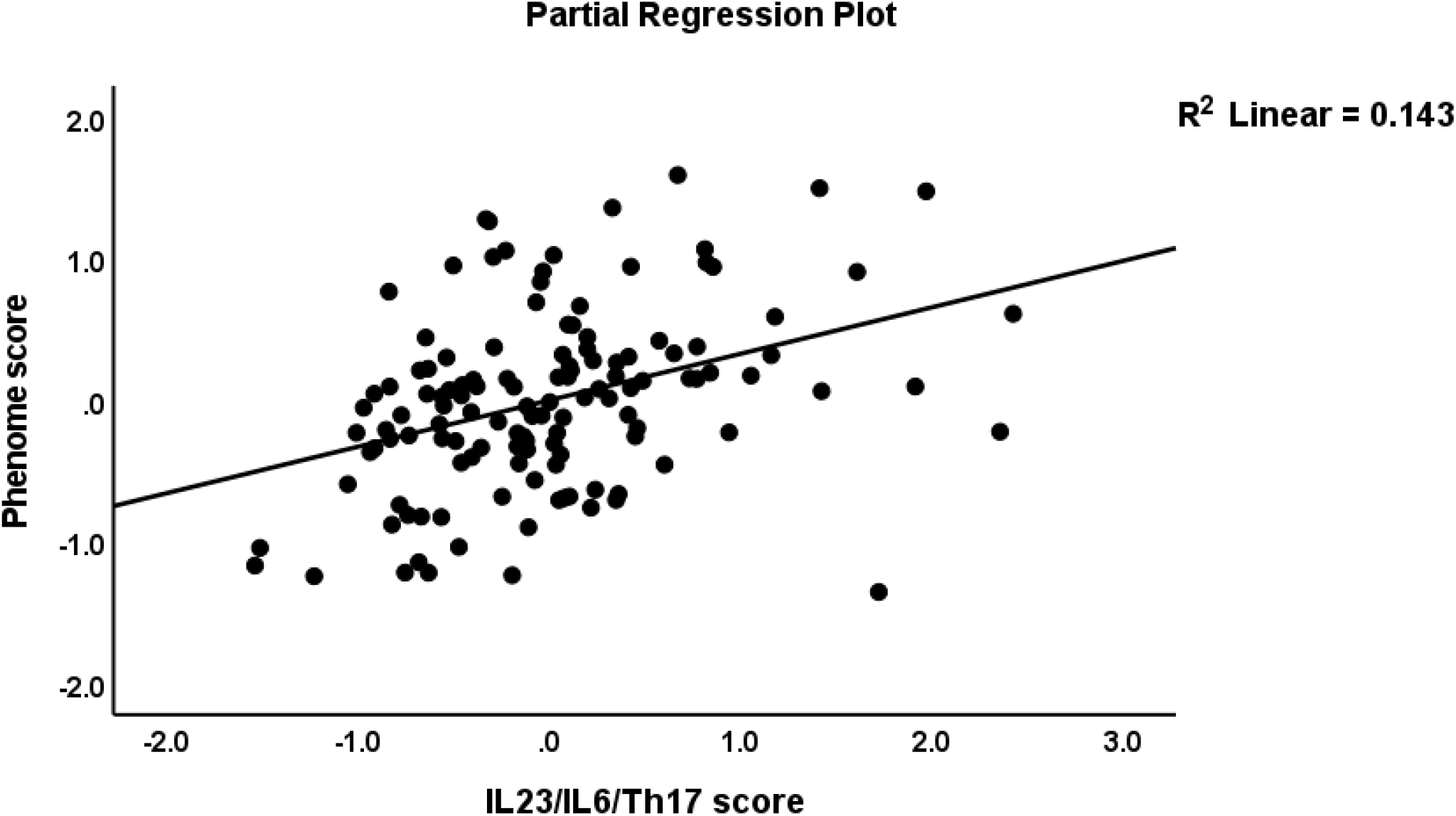
Partial regression plot of the phenome of schizophrenia on the interleukin (IL)23/IL6/T helper (Th)17 axis score

Table 4, regression #3 demonstrates that 54.6% of the variance in the G-CoDE was explained by the regression on IL6/IL23/Th17 (inversely) and magnesium, copper, and zinc (all positively). **Figure 4** shows the partial regression plot of the G-CoDE on the IL6/IL23/Th17 score (after considering the effects of the other covariates). In the restricted study sample of schizophrenia patients (regression #4) we found that the IL6/IL23/Th17 score (inversely) and magnesium and cacium (positively) explained 28.5% of the variance in the G-CoDe. **Figure 5** shows the partial regression of the G-CoDe on magnesium (after adjusting for the other variables in this regression).

**Figure 4.**
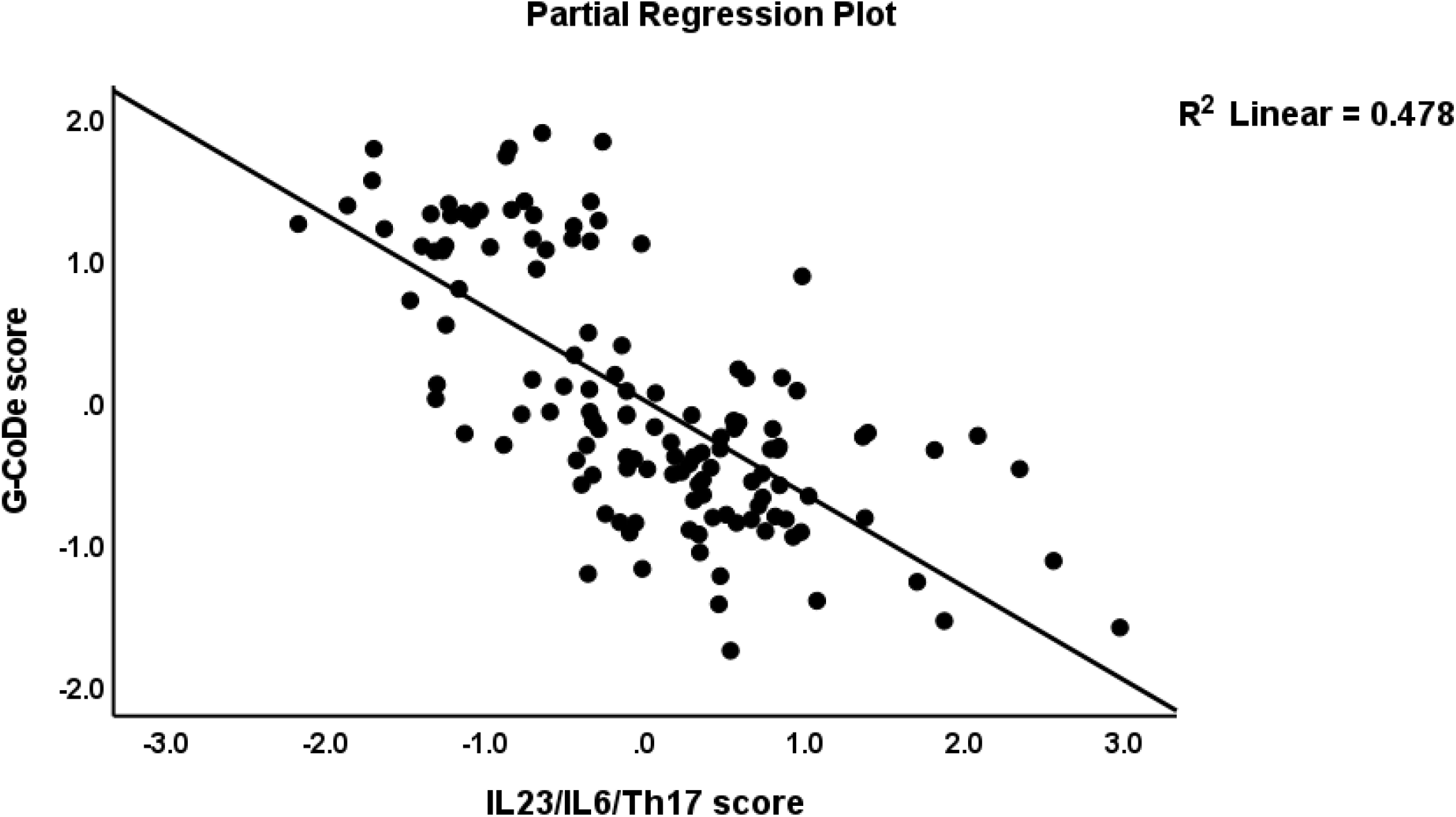
Partial regression plot of the generalized cognitive deteroriation (G-CoDe) score on the interleukin (IL)23/IL6/T helper (Th)17 axis score

**Figure 5.**
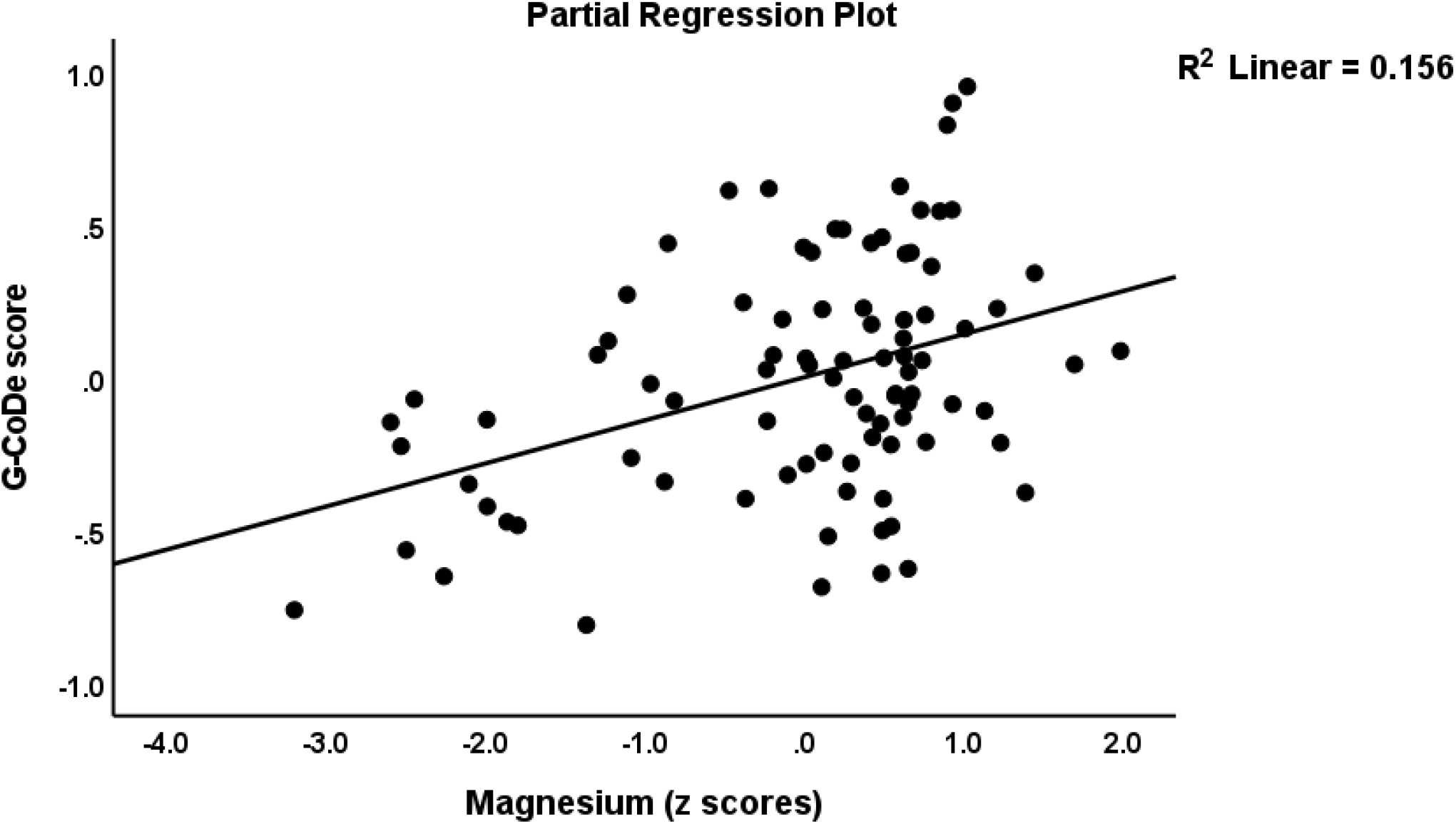
Partial regression plot of the generalized cognitive deteroriation (G-CoDe) score on serum magnesium concentrations

## Discussion

### Increased cytokine levels in MNP

The first major finding of this study is that IL-23, IL-6, IL-17, and TNF-α were considerably greater in MNP than in SNP, while IL-1β, IL-22, G-CSF, IL-21, IL-17, IL-10, and TNF-α were significantly higher in schizophrenia than in controls. Furthermore, elevated IL-23 and IL-6 were identified in MNP but not SNP as compared to controls, suggesting that these cytokines are characteristics of MNP rather than schizophrenia in general. According to previous research, MNP is associated with increased production of TNF-α, IL-6, and IL-1β, as well as CCL11, CCL2, and the soluble IL-1 receptor antagonist (59-61) and increased TNF-α levels were discovered to be a crucial factor in the pathophysiology of MNP (62).

Increments in cytokines such as IL-1β, TNF-α, IL-6, and IL-10 have frequently been described in schizophrenia (63). Blood IL-23 levels were shown to be elevated in treated schizophrenia patients, FEP and recurrent episodes of schizophrenia (29, 64-66). As reviewed by Roomruangwon et al. (63) and reported in some (28, 29, 65, 67, 68), but not all (69), studies, schizophrenia is accompanied by increased IL-17 levels. Increased IL-22 and IL-6, but not IL-17 or IL-1β, were observed in schizophrenia as compared with healthy subjects (70). Interestingly, antipsychotic treatment for 3 months was found to significantly reduce plasma levels of Th17-pathway related cytokines like IL-6 and IL-17A (71). Contradictorily, a four-week treatment with antipsychotic agents did not affect circulating levels of IL-23 or IL-17 (72,73). The differences noted above are most likely due to the comparison of various research groups, such as MNP vs. SNP/controls or schizophrenia vs. controls.

All in all, our results show that schizophenia is accompanied by indicants of activation of the M1 (IL-1β, IL-6, TNF-α) and Th-17 (IL-17, IL-21, IL-22, and TNF-α) phenotypes, and that MNP is additionally characterized by increased IL-6 and IL-23, while increments in those cytokines contribute to increased G-CSF and IL-10 (73-75).

### IL-23 and IL-6 as drivers of Th-17 pathway activation

The second major finding is that our PLS analysis revealed that increasing IL-23 and IL-6 levels are linked to Th-17 activation, as measured by extracting a latent vector from IL-17, IL-21, IL-22, and TNF-α. As described in the introduction, IL-23 and IL-6 are key factors in the growth and survival of the pathogenic Th-17 phenotype (35-38, 76, 77). Moreover, IL-6 and IL-23 signaling is required to activate STAT3 and IL-17 production, while IL-21 may further activate STAT3 (78, 79). IL-23 is a key cytokine involved in a variety of inflammatory and autoimmune disorders by producing a highly pathogenic T cell population (80). Th-17 cells are the main producers of IL-22 (81) and IL-17 coupled with IL-22 mark a particularly pathogenic population of aggressive autoreactive and pro-inflammatotory T cells (82). Nevertheless, IL-22 also has protective properties (83) including acting as an antimicrobial peptide and collaborating with IL-17 in the regulation of the immune response (84, 85). IL-21 is another pro-inflammatory cytokine that mediates antibody class switching and production, and this cytokine has pathogenic effects in (auto)immune disorders, including Sjogren’s syndrome, systemic lupus erythematosus, and psoriasis (86-88).

Thus, instead of calling this axis the TNF/IL-23/IL-17 axis (89) or the IL-23/IL-17/G-CSF axis (75) it is more adequate to denote this association as the pathogenic IL-6/IL-23/IL-21/IL-17/IL-22/TNF-α or shortened the IL6/IL23/Th17-axis. In this respect, we were able to extract one common core from IL-23 and IL-6, as well as the Th17 cytokines, confirming the concept.

It’s worth noting that in schizophrenia, activation of the pathogenic IL6/IL23/Th17 axis is linked to other important pathways. First, BDNF is adversely related with IL-23 and illness severity ratings in schizophrenia (90), and a first protein-protein interaction (PPI) network created using FEP/FES genes shows that BDNF is part of the same immune PPI network as STAT3, IL-6, TNF-α, IL-1β, and IL-10 (91). Second, complement component factors that have a role in schizophrenia, such as C1q, C3, and C4 (22, 92, 93), may affect the production of IL-23 and IL-17 family members (94). Third, BBB endothelial cells express IL-17 and IL-22 receptors, and binding to IL-17 and IL-22 causes BBB permeabilization (95, 96), thereby aggravating the effects of LPS, other cytokines and tryptophan catabolites on BBB breakdown (97). Fourth, the IL6/IL23/Th17 axis plays an important role in the immune-inflammatory responses directed to gut commensal microbiota (including Gram-negative bacteria). Thus, commensal bacteria may prime IL-23 expression and production by dendritic cells (98), and induce IL-22 production in the gut (99). IL-23 increases gut permeability by breakdown of the tight juction barrier (100) and orchestrates the immune-inflammatory response in the gut (101). IL-22 has generally more regenerative and protective functions (102-104), including the regulation of antimicrobiol peptide production and the composition of gut commensal bacteria (99). IL-17 plays a key role in the response to infectious agents by mounting an innate immune response in epithelial cells which may lead to systemic inflammatory and autoimmune responses (105). IL-21 may trigger gut inflammation and plays a role in inflammatory bowel disease, while claudin-5 is a downstream gene of this cytokine (106). Breakdown of the tight junctions and adherents barriers (leaky gut) with increased bacterial translocation is involved in schizophrenia and, especially, in MNP (97, 107).

### Effects of the pathogenic IL6/IL23/Th17 axis on G-CoDe and the phenome of schizophrenia

The third major finding of this study is that the latent vector score extracted from the IL6/IL23/Th17 data was considerably greater in MNP than SNP, with a difference of more than one standard deviation, and predicted cognitive deficits and the severity of the phenotype of schizophrenia. Moreover, we showed that a reliable latent vector could be extracted from the IL6/IL23/Th17, G-CoDe and phenome scores, thereby shaping a new pathway phenotype reflecting the link among peripheral adverse outcome pathways and the cognitome and phenome of schizophrenia. Furthermore, clustering analysis revealed two qualitatively distinct and well-defined patient groups using the same three scores, and these groups perfectly matched the deficit *vs* non-deficient dichotomy. These findings support the view that MNP and SNP are qualitatively distinct classes (108) an that the pathogenic IL6/IL23/Th17 axis is a major player in this regard.

We previously addressed IL-6 and TNF-’s many neurotoxic actions, which, when coupled with other neurotoxic substances like as LPS and lipid peroxidation, may produce neurotoxicity and, hence, explain the cognitive deficits and phenome of schizophrenia (109, 110). As discussed previously, these compounds may induce a plethora of neurotoxic effects, especially on processes like neurogenesis, neuroplasticity, cerebral cortex radial glia guided migration, synapse assembly, axogenesis, axonal spreading and branching, synapse assembly and structure, pre- and post-synaptic neuronal connectivity, regulation of excitatory synaptic functions and post-synaptic protein assembly (111). By inference, the strong effects of the IL6/IL23/Th17 axis on both the cognitive detrioration and symptomatome of schizophrenia indicate that also IL-23, Th17, IL-22 and IL-21 may further contribute to the neurotoxic effects.

A variety of cells in the central nervous system (CNS) produce IL-17 under physiological conditions, while IL-17R is expressed by immune cells and glial cell populations (112). IL-23 is produced by CNS cells and is a principal mediator of microglial activation, neuroinflammation, and tissue damage (113). There are two ways in which Th17 cells are involved in neuroinflammation, either directly via the production of cytokines (IL-17, IL-21; IL-22; IFN-γ; and G-CSF) or indirectly via the stimulation of neutrophil infiltration and microglial cells, which secrete cytokines, and by attracting CD8+ and Th1 cells to the CNS (114). IL-17 has been hypothesized to adversely influence adult hippocampus neurogenesis even in physiological conditions (115). Moreover, IL-23-activated Th17 cells in the brain are essential for sustaining persistent neuro-inflammation during infection and autoimmune responses (116). Furthermore, following stroke, IL-21 and its receptor are expressed in the CNS via brain infiltrating CD4+ T cells and CNS IL-21 strongly contributes to CNS tissue damage (117). IL-22 is expressed in the brain in physiological conditions, and is upregulated during neuroinflammatory conditions (118, 119). While this cytokine may have protective effects (119, 120) and induces an acute phase response (121) it may also display more detrimental inflammatory effects (99). Most importantly, during inflammatory responses IL-22 is upregulated in the brain and may increase the production of TNF-α, IL-6, and prostaglandins and induce STAT3, MAP-kinase, and JAK-STAT pathways (118), which all play a role in schizophrenia (118, 122).

It is also important to note that after considering the detrimental effects of the IL6/IL23/Th17 axis on the phenome of schizophrenia, IL-10 showed an inverse association with the phenome. Since IL-10 levels were higher in patients than in controls and since IL-10 is a negative immunoregulatory cytokine (20), our results indicate that a relative shortage in IL-10 CIRS functions may aggravate the effects of the pathogenic IL6/IL23/Th17 axis.

### Calcium, magnesium and zinc in schizophrenia

This study shows that schizophrenia is accompanied by lowered zinc as compared with controls, and that MNP is characterized by lowered magnesium and calcium but higher copper as compared with SNP. As described in the Introduction, some (44, 123-129) but not all (130, 131) authors reported changes in these trace elements and ions in plasma, serum or CSF in schizophrenia patients. Although some studies reported inverse associations between calcium and magnesium levels and severity of schizophrenia others could not find any association (123, 132). Nevertheless, we detected that magesium, zinc, copper, and calcium were positively associated with the G-CoDe indicating that those trace elements and ions protect cognitive functions against the neurotoxic effects of the IL6/IL23/Th17 axis and thus that the lowered levels in schizophrenia and MNP contribute to the overall cognitive deteroration.

It is important to note that inflammation or the acute phase response is accompanied by lowered levels of zinc, magnesium, and calcium (reviews in: (133, 134). Moreover, these three elements and copper all modulate NMDA receptor (NMDAR) functions: a) zinc and magnesium are anatagonists of the glutamatergic NMDAR (135, 136); b) increases in intracellular calcium exert a negative feedback on NMDA channels (137); and c) copper modulates the affinity of hippocampal NMDARs for the co-agonist glycine pocampus (138). As such, changes in the equilibrium between these trace elements and ions may be involved in the regulation of the NMDAR and NMDAR-mediated neuroplasticity and excitotoxicity and, therefore, may play a role in schizophrenia.

### Conclusion

All in all, the pathogenic IL6/IL23/Th17 axis, which may be induced by microbiota, contributes to neurocognitive deficits and the phenome of schizophrenia and especially MNP, due to its key role in peripheral inflammation, gut and BBB permeability, neuroinflammation and ensuing neurotoxic effects on CNS circuits. Such effects will appear or be more prominent in subjects with CIRS deficits including lowered IL-10, magnesium, calcium, and zinc (this study), and deficits in antioxidant, miRNA and neurotrophic defenses including in brain-derived neurotrophic factor, neurotrophin/Trk, RTK and Wnt/catenin signaling (22).

## Data Availability

The dataset generated during and/or analyzed during the current study will be available from MM upon reasonable request and once the authors have fully exploited the dataset.

## AUTHOR DECLARATIONS

### Conflicts of Interest

The authors declare that they have no known competing financial interests or personal relationships that could have influenced the work reported in this paper.

### Funding

There was no specific funding for this specific study.

## Acknowledgments

We acknowledge the staff of Al-Hakeem General Hospital, Psychiatric Unit in Najaf Governorate, for their help in collecting samples. We also acknowledge the work of the high-skilled staff of Asia Clinical Laboratory in Najaf city for their help in the ELISA assays.

## Author’s contributions

All the contributing authors have participated in preparation of the manuscript.

## Research involving Human Participants

The study followed Iraqi and international privacy and ethics laws. The study was approved by the ethics committee (IRB) of the College of Science, University of Kufa, Iraq (82/2020), which follows the Declaration of Helsinki’s International Guideline for Human Research Protection.

## Informed consent

Before participating in this study, all participants and first-degree relatives of participants with schizophrenia gave written informed consent (legal representatives are mother, father, brother, spouse, or son).

## Notes

### Competing Interest Statement

The authors have declared no competing interest.

### Funding Statement

This study did not receive any funding

### Author Declarations

The study was approved by the ethics committee (IRB) of the College of Science, University of Kufa, Iraq (82/2020)

